# The Feasibility and Effectiveness of Digital Enabled Performance-based Incentives in Ethiopia’s Community Health Program

**DOI:** 10.1101/2025.03.12.25323854

**Authors:** Yared Kifle Gebretsadik, Alemnesh Hailemariam Mirkuzie, Gizchew Tadele Tiruneh, Girma Tadesse Biru, Getnet Alem Teklu, Hailegiorgis Abate, Tenaw Chanie, Wondosen Shiferaw, Aklilu Abera, Esubalew Sebsibe, Eyoel Mitiku, Wuleta Betemariam, Desalew Emaway

**Author notes:** Corresponding author: Yared Kifle Gebretsadik. These authors contributed equally to this work (Yared Kifle Gebretsadik, Alemnesh Hailemariam Mirkuzie). These authors contributed equally to this work (Gizchew Tadele Tiruneh, Girma Tadesse Biru, Getnet Alem Teklu, Wuleta Betemariam).

## Abstract

Ethiopia is among the countries with a well-established community health program also known as a health extension program (HEP) aiming at improving access to basic healthcare services at the community level. The program has achieved remarkable success, significantly improving healthcare access, reproductive, maternal, newborn, and child health (RMNCH) outcomes, and advancing Ethiopia’s progress toward meeting national and global health targets. Despite its success, a national HEP assessment conducted in 2020 showed a decline in health extension workers’ performance, low motivation, and gaps in performance management have challenged the program. To enhance motivation, performance, and accountability, and improve service quality, the Ministry of Health integrated Performance-Based Incentive (PBI) intervention into the Electronic Community Health Information System (eCHIS). This implementation research (IR) explored the feasibility, acceptability, and adoption of digitally enabled Performance-Based incentives (PBI).

Implementation research (IR), which employed a mixed method design guided by the Reach, Effectiveness, Adoption, Implementation, and Maintenance (RE-AIM) framework [1]was conducted in three implementations and four control Woredas. The qualitative data collection utilized a semi-structured interview and discussion guides to collect data using focus group discussion, key informant interviews, and small group discussions. Content analysis was used to underpin the qualitative components of the study, and was guided by the Consolidated Criteria for Reporting Qualitative Research (COREQ) checklist. Thematic analysis was used to analyze the qualitative data. Quantitative data was obtained from eCHIS and DHIS2 dashboards and interrupted time-series analysis and paired t-tests to evaluate changes in maternal and child health services and describe performance of HEWs.

The PBI intervention was fully adopted across the intervention Woredas. The HEWs extensively utilized eCHIS for service provision, tracking their performances, reporting, and referrals. Supervisors on the other hand used the enhanced focal person application to set HEWs performance targets, provide supervisory and mentorship support, and monitor the progress of HEWs towards set targets. Adoption of the PBI was facilitated by its alignment with the existing workflow, and intergradation into the routine HEP operations. High acceptability of the intervention was reported due to its transparency, efficiency, and ability to streamline workflows. Digital tools minimized manual tasks and enhanced trust in performance evaluations and efforts to incentivize the best performers. The intervention fostered motivation among HEWs, witnessed by significantly improved KPI scores between the first and second incentive rounds (from 66 to 82, p<0.05). Supervisors and Woreda managers emphasized the intervention’s capacity to drive data-driven decision-making and performance monitoring. Feasibility was demonstrated by availing of essential digital tools (tablets, Wi-Fi routers, power banks, and fingerprint scanners), eCHIS capacity-building training, and alignment of strategies with routine health system practices. HEWs reported that the intervention simplified the tracking of performances and target-setting. However, infrastructure limitations, inconsistent mentorship, and high staff turnover were identified barriers to optimal implementation.

The digitally enabled PBI intervention proved to be feasible, acceptable, and effective in improving the health extension workers’ performance and improving RMNCH outcomes. Addressing the issue of infrastructure limitations, availing resources needed for consistently implementing PBI, and institutionalizing the intervention within the routine HEP activities remained crucial.

## Introduction

Ethiopia is among the countries with a well-established community health program also called the Health Extension Program (HEP) that aims to improve reproductive, maternal, newborn, and child health (RMNCH) outcomes [1]. The Health Extension Program (HEP) is one of the innovative community-based health programs launched in 2003 by the Ministry of Health (MoH) to make health services accessible to rural communities, the HEP was designed to provide promotive, preventive, and curative primary health services to rural communities by setting out Health Extension Workers (HEWs) in rural Health Posts (HPs). The HEWs are trained, salaried female community health workers, who act as the primary implementers of the HEP. Over the last decade and a half, HEP has proven to be an effective intervention by serving as the largest component of Ethiopia’s health care delivery system in terms of reach and thus transforming access to health care services. Moreover, the HEP has made a solid contribution to Ethiopia’s recent gains in health outcomes in terms of increasing demands for health services and decreasing morbidity and mortality related to nutrition, communicable diseases, and maternal and child health. The HEP is still considered to be one of the major drivers for achieving national as well as global health targets and attaining universal health coverage (UHC) [2], [3]

Although the HEP is well acknowledged for its remarkable contribution to improving the RMNCH outcomes and achieving Universal Health Coverage (UHC), the national HEP assessment conducted by the Ministry of Health (MoH) in 2019 revealed that the program has experienced a decline and loss its momentum due to several challenges. One of the significant challenges identified was the absence of structured and systematic motivation packages for the HEWs which led to poor motivation and declining performance among HEWs and their supervisors [4]. The report also highlighted a critical knowledge gap in understanding how to systematically and effectively improve HEW performance and motivation. Although global evidence suggests that performance-based incentives (PBIs) are a promising approach to enhancing workforce performance and accountability [5], there is limited knowledge about how such systems can be integrated and scaled within Ethiopia’s community health system framework in general and eCHIS in particular. Moreover, questions regarding the feasibility, acceptability, and sustainability of PBIs in a digital health framework, such as the Electronic Community Health Information System (eCHIS), remain unanswered. Recognizing these challenges, the MoH proposed Performance-Based Incentives (PBIs) as a key strategy in the HEP Optimization Roadmap (2020–2035) to enhance service access, quality, and HEW performance [4]. PBI connects measurable indicators to rewards, promoting accountability and aligning individual goals with broader health outcomes. The relationship between PBI and workforce motivation, health worker performance, service provision, and quality are well documented [6], [7].

The MoH in collaboration with JSI designed and tested the digitally enabled PBI innovation to test the impact of eCHIS enabled PBI system on the HEW’s performance. The study provides valuable insights into how digital tools can enhance HEW motivation, service quality, and data-driven decision-making. The findings demonstrate significant improvements in HEW performance, RMNCH service coverage, and the use of data for decision-making.

## Methods

### Study settings

The Ethiopian primary healthcare structure in a Woreda (i.e., district) health system is organized around a primary hospital, 4-5 Primary Health Care Units (PHCU) composed of health center (HC), and 5 cluster health posts (HPs) serving a catchment population of approximately 25,000. Over 17,500 HPs in the country and over 40,000 Health Extension Workers (HEWs) provide essential health services through the program [4], [8].

Since its launching in 2003, the HEP has remained an integral part of the primary and flagship community-based healthcare delivery system in the country [9]. The HEWs are trained and salaried community health workers providing health promotion, disease prevention, and curative primary health care services [8]. The HEP used a paper-based health information system until 2018 when the MoH spearheaded the transition to an electronic community health information system (eCHIS). By 2024, the eCHIS has been expanded to nearly 8,000 health posts in the country [10]. eCHIS, the tablet-based, community-centered information system not only digitizes health data but also acts as a job aid for HEWs. It supports high-quality care by providing prompts for treatment protocol adherence, appointment reminders, consistent follow-ups, and referral linkages to the next level of health facilities such as cluster health centers.

By capturing data in real-time at the point of care, eCHIS automates the sharing and reporting of critical health indicators, enhancing the overall efficiency of health service delivery. Three applications support eCHIS: the HEW Application assists HEWs in managing Family Folders for service delivery and follow-up, with each HEW having their devices equipped with this mobile application. The HC Referral Application enables health center workers to confirm referrals and give feedback to HEWs about those referrals. The Focal Person Application (FPA) assists health center focal persons in providing technical and programmatic support to HEWs. This project has enhanced the existing FPA within the eCHIS framework. The FPA includes features such as the Plan Setting Manager, which allows for the setting of the HP’s eligible population and targets at the beginning of each fiscal year, and the Supervisor Task Manager, which supports supervisors in their routine activities by integrating supportive supervision and mentorship checklists. This functionality enables supervisors to oversee tasks and challenges identified during mentorship and supervision at the HPs. The Performance Management (PM) Dashboard allows them to monitor the performance of HEWs and HPs, manage activities, generate reports, and facilitate data-driven decision-making to improve accountability and efficiency (Fig 1).

**Fig 1:**
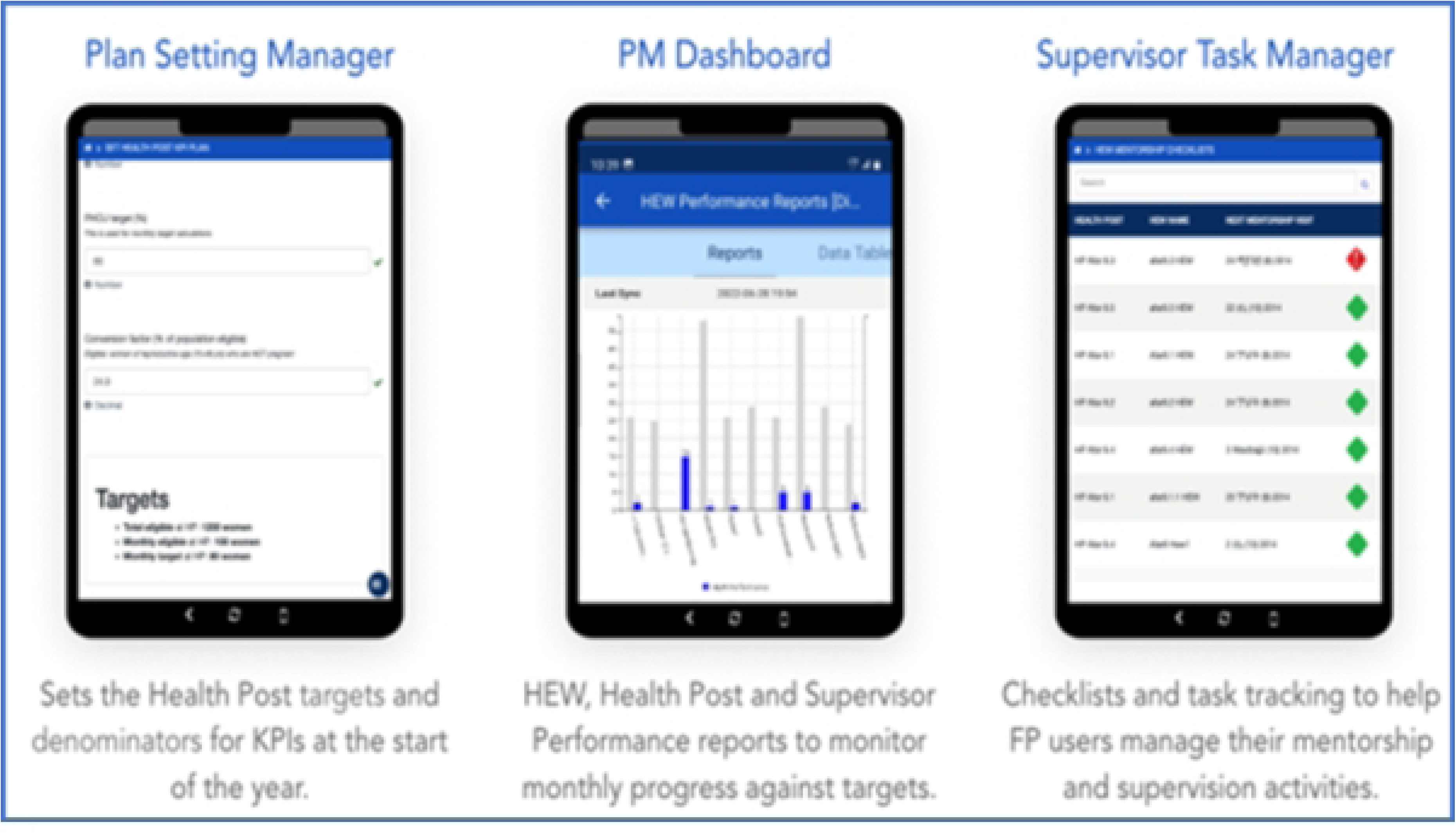
Enhanced focal person application features.

JSI has been supporting the MoH’s eCHIS scale-up initiative through funding from the Children’s Investment Fund Foundation (CIFF) since 2020 as part of a five-year project. By November 2024, JSI had facilitated the eCHIS scale-up in 2,135 health posts across 110 Woredas. In July 2023, JSI has supported the MOH in designing and testing digitally enabled PBI on the eCHIS platform for improving HEWs performance, service quality, data use for decision, and improving maternal and child health outcomes. This was followed by selection of three implementation Woredas-Awabel in the Amhara region, Gimbichu in the Oromia region, and Mirab Abaya in the Southern Ethiopia region and four control woredas-Arbaminch zuria from South region, Liban Chukala from Oromia, Damot Pulassa, and East Badawacho form Centeral Ethiopia region. These Woredas were chosen in collaboration with Regional Health Bureaus (RHBs), local stakeholders, and other partners to ensure the intervention was context-specific and appropriate.

### Descriptions of the interventions and implementation strategies

The design of PBI interventions was guided by the Digitized, Equipped, Supervised, and Compensated (DESC) framework considering operational aspects, including digital enablement, equipping HEWs, supervision, and compensation [7], [11]. The PBI intervention utilized enhanced features of the eCHIS platform, including the HEWs’ app, Focal Person Application, and national eCHIS dashboard. (Fig 1) Key intervention components included:

1. **Digitally Enabled Tools**

- HEWs utilized eCHIS for service delivery, data collection, and reporting.
- Supervisors leveraged FPA for target setting, performance tracking, and supervision.
- Real-time data dashboards facilitated decision-making.
2. **Equipping for the intervention**

- Training for HEWs, supervisors, and managers. Provision of essential drugs, medical equipment, high-spec tablets, power bank, and internet connectivity.
3. **Supervisory Support**

- Integrated mentorship and supervision through digitized checklists.
4. **Compensation Mechanism**

- PBI was awarded bi-annually (in two rounds) based on KPI scores for HEWs, HPs, supervisors/health centers in the top 1-3 ranks (Key Performance Indicator (KPI): KPIs are measurable values that help track the progress of PBI implementation in achieving set targets. In partnership with the Ministry of Health (MOH), we established 22 KPIs—10 for HEWs, 6 for HPs, and 6 for HC supervisors. These KPIs were used to monitor implementation progress in real time, enabling performance quantification and playing a crucial role in incentivizing HEWs, HPs, and HC supervisors).
- In-kind incentives include laptops, Wi-Fi-routers, smartphones, trophies, and handbags.

eCHIS dashboard was used by Woreda and HC decision-makers for real-time performance monitoring, evaluation, performance verification, and handing out incentives for high-performing HEWs, supervisors, and HPs. The implementation strategies employed were target setting, training, supportive supervision, performance review, data used for monitoring, and decision and incentivization.

### Study design

This is implementation science research that employed a mixed-methods approach guided by the Reach, Effectiveness, Adoption, Implementation, and Maintenance (RE-AIM) framework [11], [12]. The quantitative component of the study used a quasi-experimental observational controlled design with three interventions and four comparison Woredas. Intervention Woredas implemented digitally enabled PBI innovations embedded within the eCHIS platform from October 2022 to November 2023. Comparison Woredas implemented eCHIS without PBI during the same period. Data were triangulated [13] from process evaluation eCHIS, DHIS2 dashboards, interviews, focus groups, and document reviews to ensure consistency between data and findings. The qualitative data were gathered through focus group discussions, in-depth interviews, and, observations conducted at baseline, process, and end-line evaluations. To guide the data collection and analysis and reporting we followed the Consolidated Criteria for Reporting Qualitative Research (COREQ) [14] framework. The quantitative component of the study used a quasi-experimental controlled observational design having three woredas implementing the PBI intervention and four comparison woredas.

### Data sources, participants, and data collection

The study compiled primary and secondary data from baseline, process, and end-line evaluations triangulating qualitative and quantitative data. The baseline data were collected in Oct 2022 and the process evaluation was conducted during review meetings between the implementation period and end-line data was gathered in Nov 2023. The primary qualitative data were collected through semi-structured interviews (n=60), in-depth interviews (n=43), and focus group discussions (n=5, 34 participants), from intervention Woredas. During the baseline phase, 24 small group discussions were conducted with 64 participants to gather initial qualitative data and inform the co-design of the intervention. In the process phase, 8 key informant interviews (KIIs) were conducted with 8 participants. In addition, 5 focus group discussions (FGDs) were held with 34 participants, and 7 small group discussions were conducted with 18 participants to explore collective experiences and gather feedback during the intervention. During the end-line phase, 4 key informant interviews were conducted with 10 participants, and six small group discussions were held with 12 participants to evaluate the outcomes of the intervention. In total, across all phases, there were 8 KIIs with 18 participants, 42 small group discussions with 94 participants, and 5 FGDs with 34 participants.

Qualitative data collection used a pretested interview guide with RE-AIM dimensions as the main theme and probing questions. We used FGD and in-depth interview guides specific to the different categories of informants (HEWs, supervisors, Woreda, and HC experts). The guides prepared in English were then translated into local languages (Amharic and Afan Oromo) and pretested before use. Experienced data collectors trained on the study objectives, interview guides, and ethical principles collected the data. Before the interviews and FGDs, the objectives, risks, benefits, and voluntary participation of the study were explained to the participants. Informed written consent was obtained during end-term evaluation and verbal consents were obtained during review meetings. Respondents’ confidentiality, privacy, and anonymity were respected throughout the process. Audio recordings of the conversations were upon the informants’ permission. Informants were purposively selected to capture diverse experiences and perspectives.

The Key informant, in-depth interviews, and FGDs were conducted face-to-face in the informants’ natural settings, i.e., their workplaces and during review meetings [15]. Recognizing that data saturation entails the richness of the data rather than just the number of informants, we actively monitor the data saturation throughout the data collection [16]. At the end of the interviews and FGDs, the data collector undertook member checking by summarizing the main points of the discussions and sought approval from the informants. For the FGDs, the moderator leads the discussion while the note taker takes notes, quotes, and audio-recorded the dialogue. The interviews with HEWs and key informants lasted an average of 30 minutes, while the FGDs lasted 60 minutes on average per group. The secondary quantitative data were accessed from eCHIS and DHIS2 platforms for the 87 interventions and the 180 comparison health posts between October 2022 and November 2023.

### Measurement

The implementation outcome measures and data sources are presented according to the RE-AIM dimensions below (Table 1).

**Table 1:** RE-AIM dimensions, outcome measures, data sources, and analysis techniques.

| RE-AIM dimensions | Measures | Data sources |
| --- | --- | --- |
| **Reach**<br>The proportion of the eligible population receives eCHIS-based service delivery | • Number of HPs, HEW, HC, and supervisors participated in digitally enabled PBI interventions.<br>• Percentage of households, and registered on eCHIS;<br>• Percentage of HPs providing maternal and child health services using eCHIS<br>• Number of HEWs and Supervisors and woreda experts who received PBI training<br>• Number of HPs set performance target for KPIs<br>• Number of HEWs, supervisors, and HPs incentivized<br>• Number of HHs and population reached<br>• Number of clients received service using the eCHIS platform | eCHIS dashboard and CommCare database |
| **Effectiveness**<br>Effectiveness refers to the impact of the PBI strategy on the digitally enabled service delivery, motivation, and performance of the HEWs. | • Percentage of maternal and child health services provided on eCHIS<br>• Change in delivery and uptake of maternal and child health outcomes<br>• Improved motivation of HEWs and their supervisors<br>• Improved frequency and content of supportive supervision<br>• Number of HPs completed HH and member registration<br>• Number of children who received BCG, measles, and Penta vaccination using eCHIS<br>• Number of women who received ANC 1, PNC, | Program data<br>eCHIS dashboard and CommCare database<br>Baseline, process, and end-line evaluation |
|  | and contraceptives using eCHIS |  |
| **Adoption**<br>Adoption refers to the intention, initial decision, or action to employ digitally enabled PBI about eCHIS. We measured adoption by assessing the degree to which HEWs implemented the PBI innovations, identifying the barriers that hindered their adoption, and determining actions needed to enhance the adoption process. | • Number of HPs, HEW, HC, and supervisors participated in digitally enabled PBI intervention.<br>• Number of HPs and supervisors setting targets digitally using FPA<br>• Support and mentorship provided using FPA<br>• Number of HEWs used eCHIS data for monitoring their progress and generating monthly reports<br>• The proportion of HPs and HCs using the eCHIS app for service provision | Program data<br>eCHIS dashboard and CommCare database<br>Process evaluation |
| **Implementation**<br>Degree of implementation of PBI and contextual factors that hinder or facilitate the delivery of the strategies. It includes the acceptability, fidelity, feasibility, and implementation barriers and facilitators. | The acceptability, feasibility, and fidelity of the digital-enabled PBI interventions, performance measurement, and incentive | eCHIS dashboard and CommCare database |
| **Acceptability**<br>Acceptability is the perception among stakeholders that PBI is agreeable/willing to accept it [17]. We defined acceptability as how well the HEWs, supervisors, and Woreda experts received the PBI intervention and implementation strategies and the extent to which the intervention or its components might meet their needs and the HEP system they operate. | Guided by the framework from Cartwright and Francis (2017) [17], we qualitatively explored four key parameters of acceptability:<br>1. Attitude, which reflects how individuals feel about the intervention;<br>2. Intervention coherence, indicating the extent to which individuals understand the intervention;<br>3. Perceived effectiveness, capturing how individuals believe the intervention can improve performance; and<br>4. Self-efficacy, representing the extent to which individuals feel willing and competent to implement the innovation [18]. | Process evaluation |
| **Feasibility**<br>Feasibility is defined as the practical implementation of digital-enabled PBI at the health post level using the eCHIS platform | The feasibility of the digital-enabled PBI interventions, performance measurement, and incentive | Process evaluation |
| **Fidelity**<br>The degree to which PBI was implemented as it was designed | Fidelity to the various elements of the intervention's key components and strategies, including consistency of delivery as intended and the time of the implementation including digital target setting, support and mentorship, use of FPA for support, performance review and evaluation, and incentivization<br>Number of HEWs, supervisors, and HPs incentivized, data use for decision<br>Number of HEWs, HCs, and Woreda experts trained on PBI | • Program data<br>• eCHIS and CommCare database<br>• Process evaluation |
| **Maintenance**<br>The extent of PBI maintained or institutionalized in a HEP/PHC healthcare system. | Local capacity created for maintaining troubleshooting and mentorship<br>Integration of PBI into the routine workstream | Program data<br>Process evaluation |

### Data analysis

Qualitative data were analyzed using thematic analysis guided by the RE-AIM framework under predefined

RE-AIM themes. Two research team members 1^st^ coded the data separately and then brought it together, and identified codes for discussion. The expanded notes were condensed, summarized, and synthesized thematically and used during the analysis. Audio records of the interviews and FGDs were transcribed and the transcript texts were analyzed manually. Ideas and statements related to the study objective were grouped under respective predefined themes as there was no other emerging theme. Clear presentation of themes enhances the credibility of the research; well-articulated themes backed by evidence are more likely to be valid and trustworthy [19].

Preliminary analysis started in the field simultaneously with data collection by summarizing the main points from the field notes and audio recordings. Later, the transcribed interview and FGDs were read, and reread to familiarize ourselves with the data through repeated readings followed by highlighting and annotating texts to identify key themes and patterns. Similar ideas were grouped to derive broader themes, illustrated using notes to show relationships. This approach facilitated deep engagement with the data, allowing for meaningful connections to the research questions. The findings were documented in a narrative format supported by quotes, reflecting the richness of the informants’ experiences. We provided context for each quotation, explaining its connection to the specific theme, thereby enhancing the credibility of the findings by grounding them in participants’ lived experiences. Furthermore, each quotation is properly attributed using a consistent identifier (participant numbers), to maintain anonymity and confidentiality, while allowing readers to trace findings back to specific respondents.

Quantitative analyses of the secondary data obtained from the eCHIS and DHIS2 platforms involved descriptive statistics, interrupted time-series analysis, and student’s t-tests to evaluate pre- and post-intervention changes. Training on PBI was given in October 2022; hence October 2022 was taken as a baseline/treatment period to compare the before and after intervention effects. The effectiveness of the study was assessed by looking into the average monthly KPIs score and RMNCH service delivery on eCHIS comparing before and after PBI intervention. Using an interrupted time-series analysis on Stata version 15 software examined the counterfactuals of the PBI interventions. Due to the active war and state of emergency in the Amhara region, there was incomplete/no data from Awabel from July to October 2023, hence the Woreda level analysis excluded the date for Awabel for the stated period. For assessing the pre- and post-test training scores student t-test analysis was used. P value <0.05 was the cut-off to declare a statistically significant difference.

### Ethical considerations

Ethical clearance for the study was obtained on June 30, 2022, from the Ethiopian Public Health Association (EPHA) Research Ethics Review Board reference number EPHA/OG/886/22. The protocol was exempted from human subject oversight through JSI Research and Training Institute, Inc. (JSI), USA. Study permits were sought from regional and zonal health departments, Woreda health offices, respective HCs, and HPs.

Study participants were informed about the purpose of the study, its benefits, risks, and their right to opt-out. Moreover, participants were given information that their participation was voluntary and the researcher kept the information obtained from them confidential and only shared it with the study team to respect confidentiality and privacy. All interviews were conducted after obtaining respondents’ informed verbal consent.

## Results

These findings are structured around key REAIM dimensions/themes, including the acceptability, feasibility, and adoption of the digitally enabled PBI intervention; changes in HEWs motivation and performance, and improvements in RMNCH outcomes.

### Reach and effectiveness of the intervention

#### Reach

The PBI intervention covered 154 HEWs, 15 supervisors, 87 HPs, 15 HCs, and three Woredas, serving an estimated 342,116. The percentage of households registered on eCHIS in the intervention and control Woreda by the end of October was 100% and 94% respectively. Before the PBI intervention, the percentage of HPs providing BCG vaccine using eCHIS during the given month was similar in the intervention and comparison Woredas. Following the intervention, the percentage of HPs providing BCG vaccine using eCHIS showed a significant increase in the intervention Woredas (t-test P<0.01), whereas a notable decrease (t-test, P<0.01) was observed in the comparison Woredas (Fig 2).

**Fig 2:**
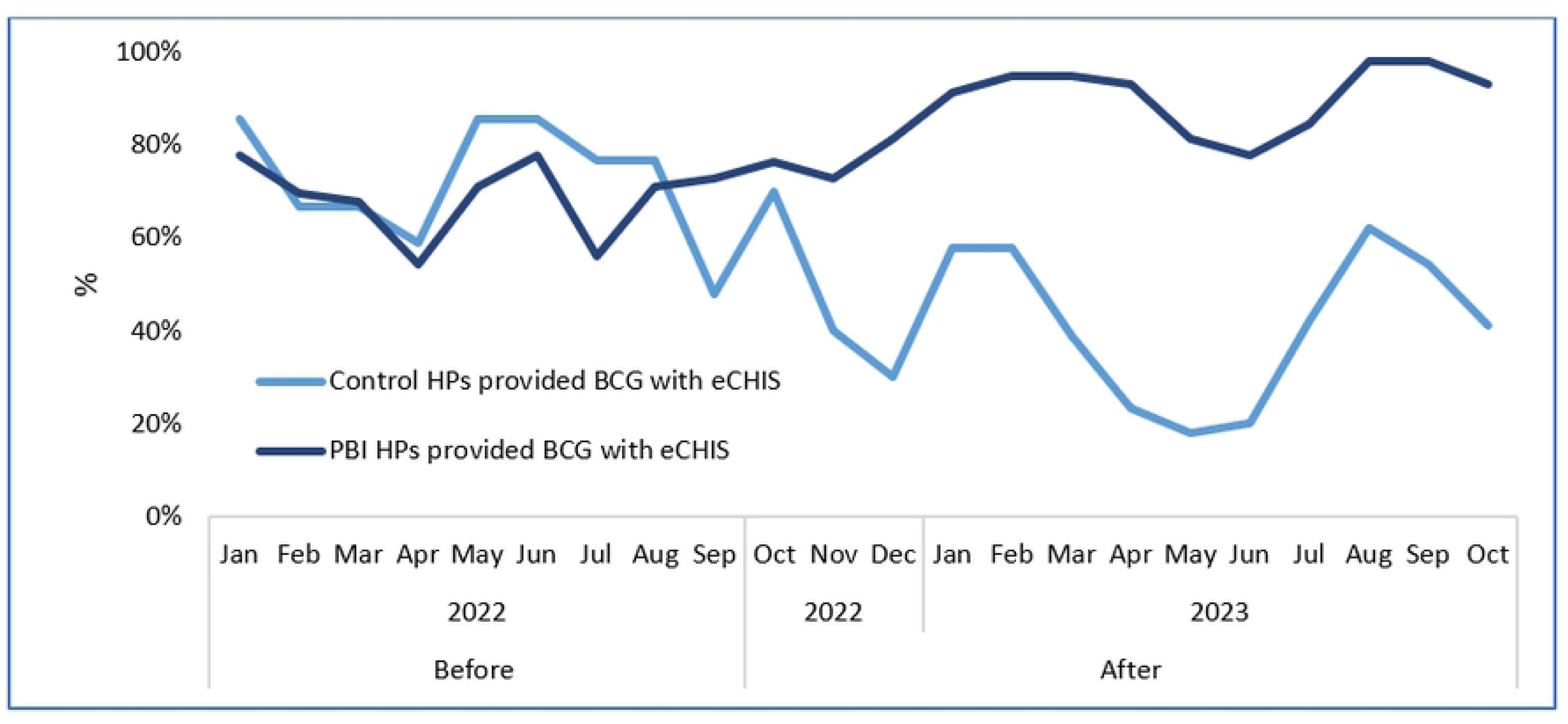
Percentage of HPs providing BCG vaccination service using eCHIS from January 2022 to October 2023 in PBI intervention and comparison Woredas.

The intervention trained 261 participants, including HEWs, supervisors, and managers. A two-tailed paired sample t-test revealed a significant improvement in the mean knowledge score, increasing from 68% in the pre-test to 75% in the post-test (p<0.001). Over 95% of trainees reported favorable reactions to the training. Supervisors set annual performance targets for all 87 health posts digitally using the Plan Setting Manager feature in the FPA. Targets were set for the KPIs from the estimated eligible catchment population by engaging respective HEWs. The KPIs with their targets were integrated into the eCHIS dashboard for effective tracking of performances and to streamline program monitoring and evaluation.

#### Effectiveness

##### A) Improved eCHIS-based RMNCH service delivery

There was a significant increase in the mean number of maternal health services provided using eCHIS following the introduction of the PBI intervention as shown in Fig 3. The mean ANC 1st visit was almost tripled (mean=155) following the implementation of PBI before (mean=56) (t-test p<0.001). By contrast, in the comparison of Woredas, the mean ANC 1st visit was reduced by half (mean=12) after the PBI intervention compared with the before (mean=27) (t-test p<0.001).

**Fig 3:**
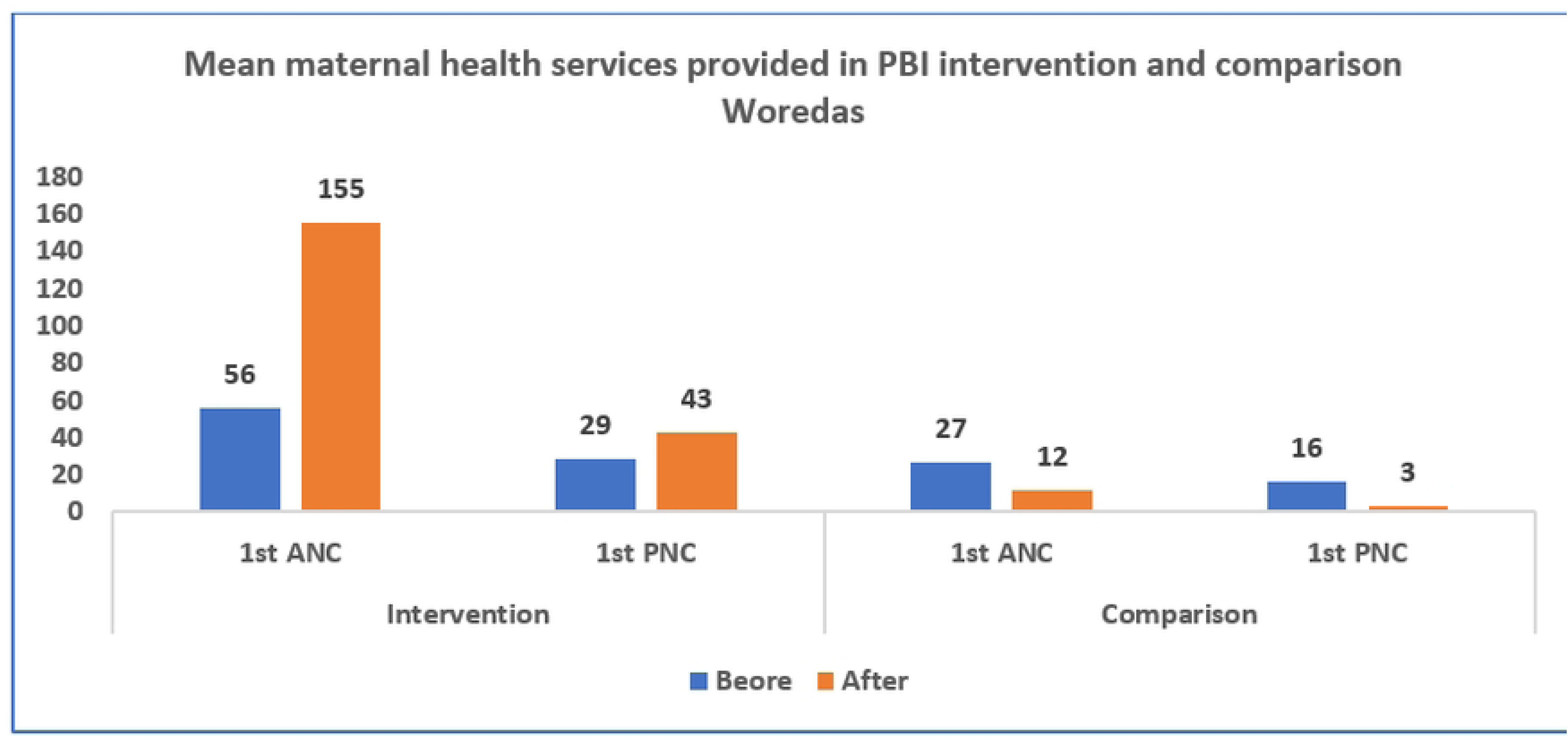
Mean maternal health services provided in PBI intervention and comparison Woredas.

The mean childhood immunization services registered on eCHIS after the intervention have shown a significant increase in PBI implementing Woredas while showing a significant decrease in comparison Woredas (t-test p<0.01). (Fig. 4)

**Fig 4:**
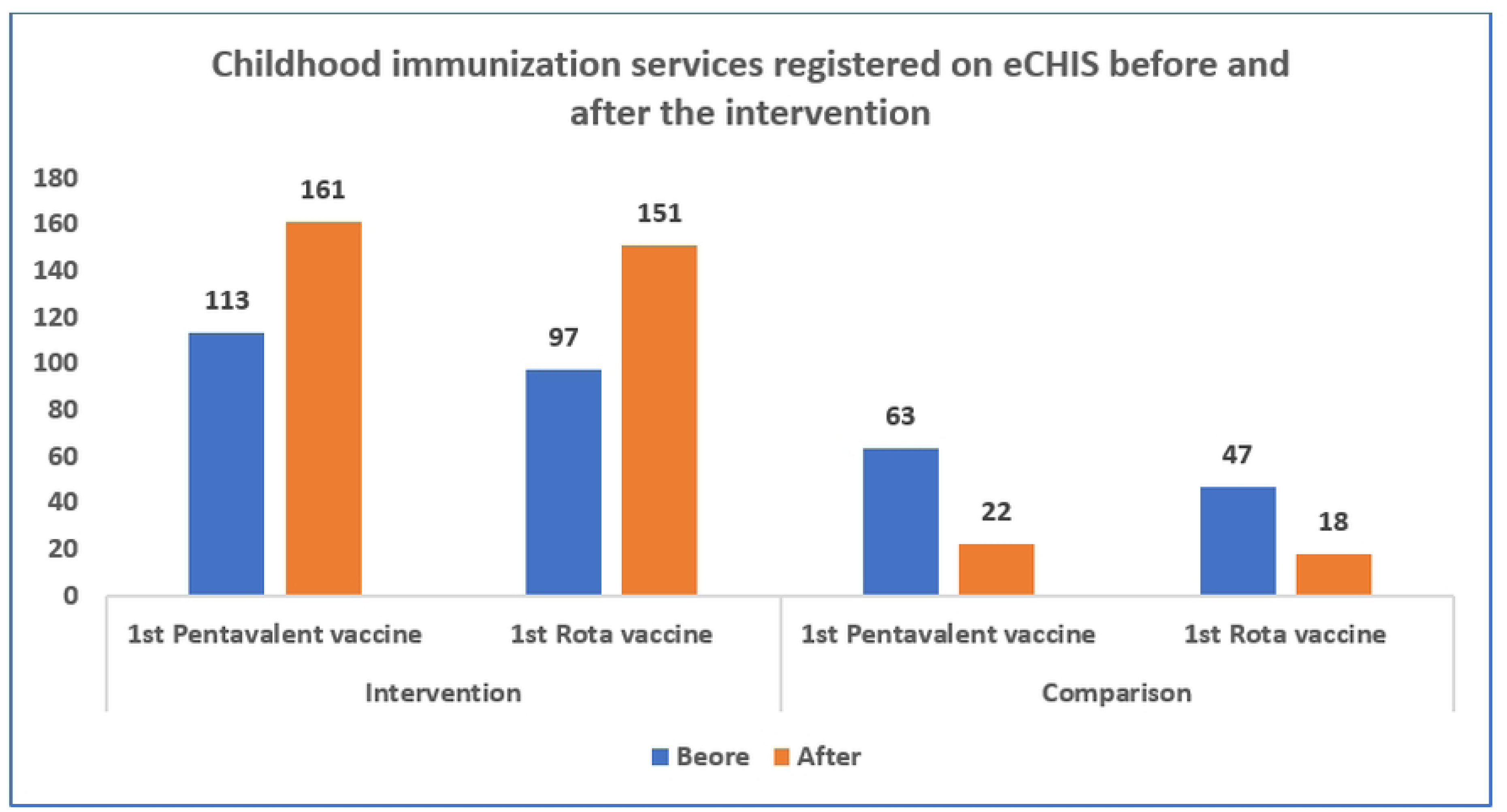
Childhood immunization services (Penta 1 and Rota virus vaccines) registered on eCHIS before and after the intervention.

##### B) Improved coverage of RMNCH-related KPIs

As shown in Fig 5, the first measles vaccination given at nine months of age registered on the eCHIS system has shown a significant consistent increase in the interventions Woreda (P< 0.01). Comparing intervention and control Woredas, there was a non-significant difference in the pre-existing levels and trends of measles vaccination in the intervention and control Woredas (P< 0.05), indicating that they were comparable at baseline. Following the intervention, the number of children who received the measles vaccine increased by 47 per month (P<0.01), with a sustained change in trend (P<0.05) indicating that the intervention drives improvement in RMNCH service provision with eCHIS.

**Fig 5:**
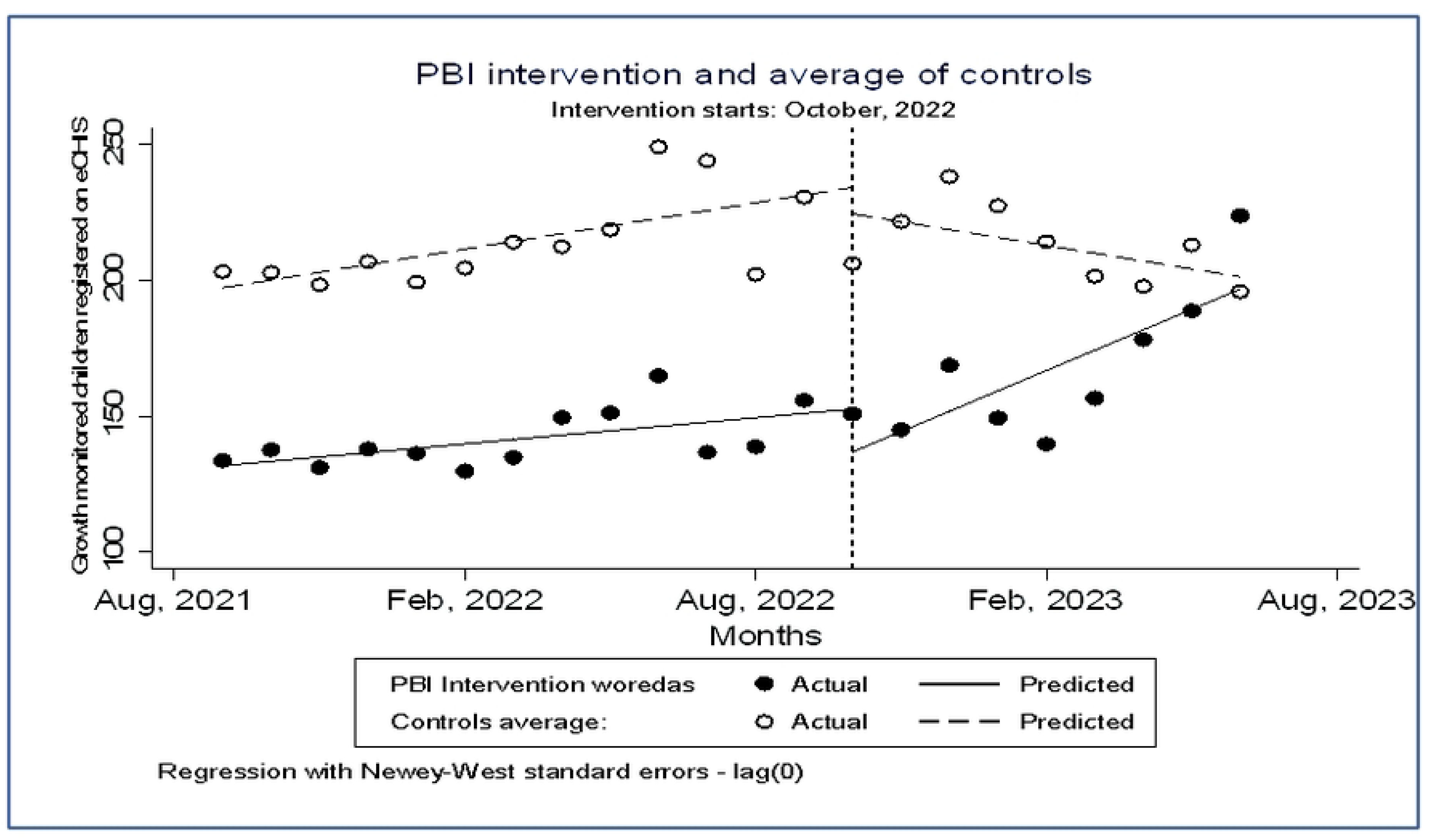
First measles vaccination uptake in PBI intervention and comparison Woredas, using data from the eCHIS platform.

Similarly, coverage of growth monitoring varied significantly according to the data from the DHIS2 platform. After the intervention, the number of children who attended the growth monitoring session in the PBI implementation Woredas showed a significantly increasing trend compared to the counterfactual (P<0.05). By contrast, there was a decreasing trend in the mean number of growths monitored children compared to the counterfactual (Fig 6) in the comparison Woredas.

**Fig 6:**
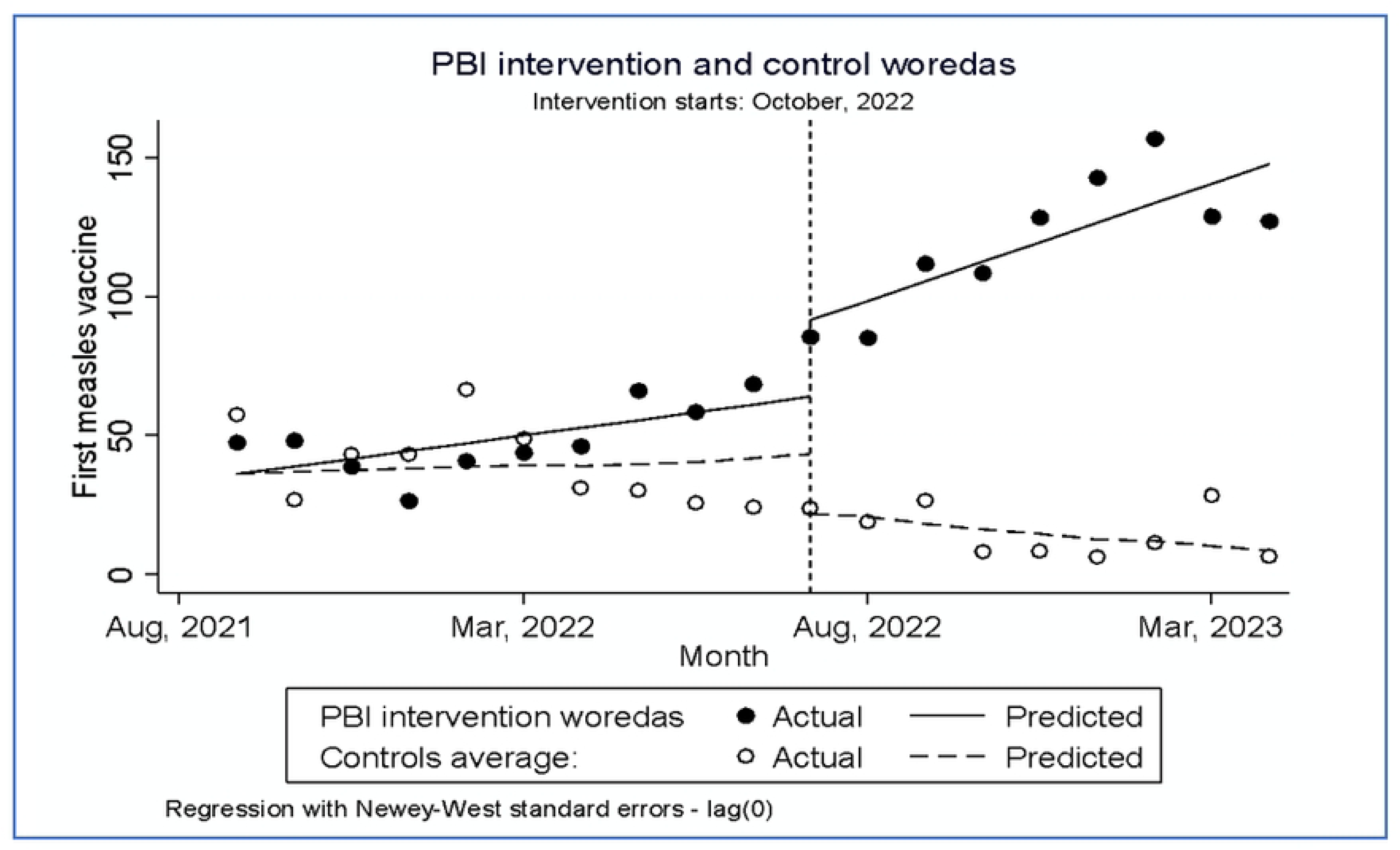
Growth monitoring performance in PBI implementation and comparison.

**Fig 7:**
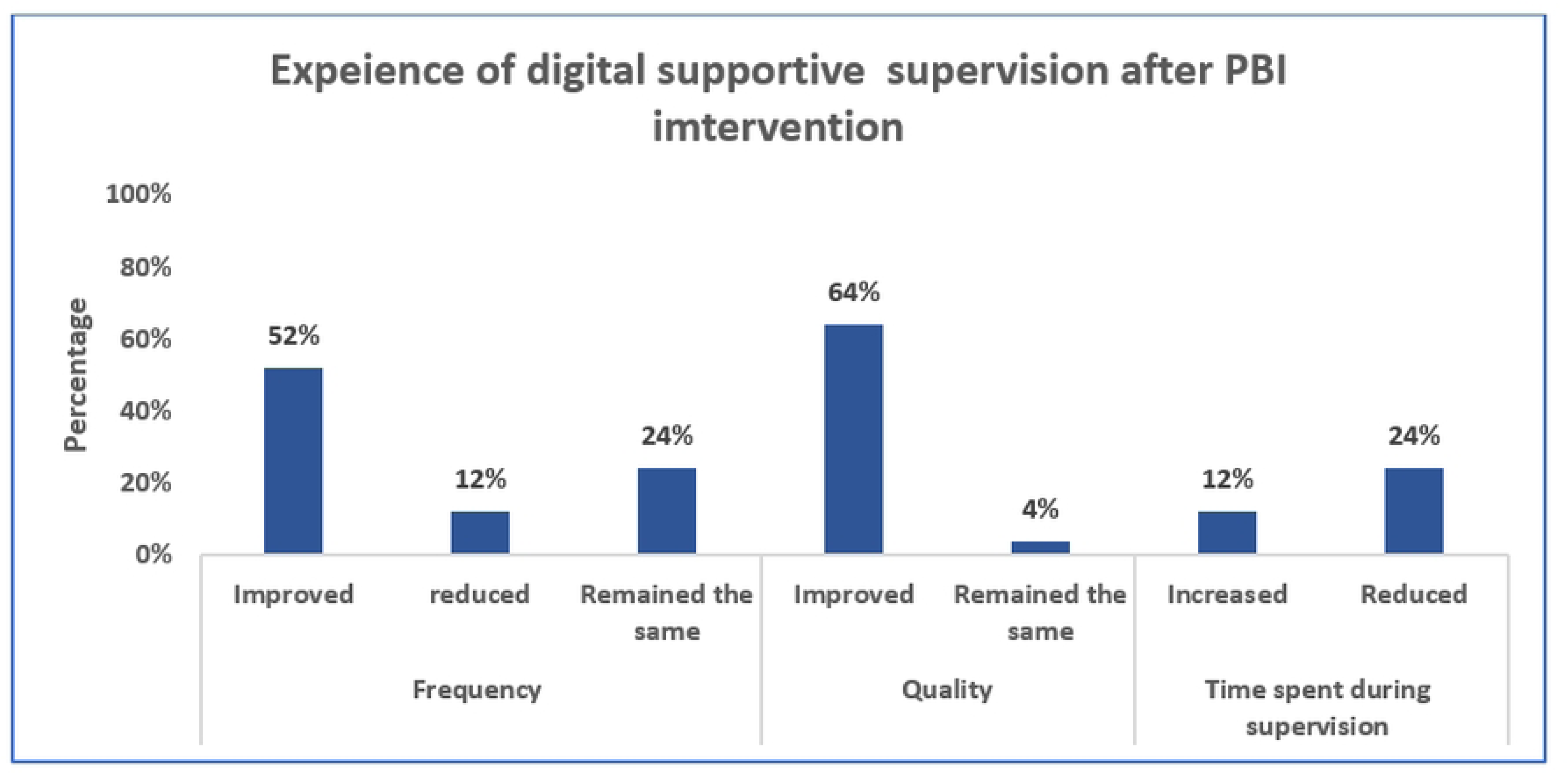
Perceived improvement in supportive supervision post digitally enabled PBI implementation.

##### C) Improved performance and motivation of HEWs

According to the data from the eCHIS platform, the KPI score during the second round of the PBI award was significantly higher than the score during the first PBI awards in all categories. For the HEWs, the average score for the 10 KPIs has increased by 16 points from 66 to 82 during the second-round award compared to the first round (t-test p <0.05). For HPs, the average score for the six KPIs has shown a 10-point significant increase during the second round compared to the first round, from 54 to 64 (t-test p <0.05). Similarly, average scores for the six supervisor KPIs have shown 17 points significant increase in the second round than in the first round i.e., from 56 to 73 (t-test p <0.01).

According to the qualitative accounts, the digitally enabled PBI intervention improved HEWs’ performance and motivation as witnessed by their supervisors, Woreda managers, and the HEWs themselves. HEW after the first award said that “I have started going house to house even after working hours to provide HEP service …”. (HEW 4). Another HEW who received an award during the second round said “The incentive has increased our motivation. After seeing the first incentive award to high-performing HEWs, we were highly motivated to perform better and hence we got awarded in the second round. (HEW 5). This was supported by a Woreda manager who said “The HEWs were highly motivated following the first incentive award. The ones who stood back were working hard to improve their rank. We have seen that practically. Digital-enabled PBI enhances performance by empowering HEWs to effectively monitor, and manage cases requiring their attention while taking appropriate feedback from their supervisors timely. “ (KI 10).

Incentivizing individual HEWs and supervisors was seen as an effective strategy for motivating them to strive for better performance. A health center director (KI 6) stated “I believe that individual incentives would further enhance motivation and performance among supervisors. It is important to continue all categories of incentives, including group and individual incentives”.

##### D) Improved supervisory support

Supervisors regularly provided support to the HEWs and HPs through digital mentorship and supervision checklists respectively to provide support. The HEWs perceived that the quality, frequency, and time spent during supervision had improved following the intervention.

Although data extracted from the eCHIS dashboard reveals improved use of the focal persona app for supervision, according to the FGD informants, FPA usage for supervision was limited due to competency gaps among supervisors to use the application attributed to inadequate training, lack of frequent practice due to competing priorities and not having the tablet in their hand, and poor motivation among some supervisors. The FGD participants stated, “FPA has been used by some but other supervisors are not well versed with the application. On the eCHIS implementation, much focus has been given to capacitating HEWs but capacitating the supervisors is equally important as they are the one who supports the HEWs. The training on the FPA was inadequate” (FGD 1)

#### Adoption of the innovation and implementation strategies

The digitally enabled PBI intervention has been successfully implemented across all PBI intervention Woredas and health posts. According to data from the end-line evaluation, informants reported that the eCHIS platform has been well adopted as the primary system for various critical functions. These include service provision, client referral, population, and household registration, tracking progress, generating monthly reports, and using data for informed decision-making at the local level (Fig 8). The platform’s ability to streamline and digitalize these processes has significantly enhanced efficiency and reliability in service delivery. Moreover, its adoption has fostered a culture of data-driven decision-making among health workers, supervisors, and Woreda health offices ensuring that resources are allocated effectively and progress is closely monitored. This comprehensive adoption not only highlights the successes of the intervention but also provides valuable insights into areas that require further refinement, offering a roadmap for future strategies to optimize the overall impact of the intervention.

**Fig 8:**
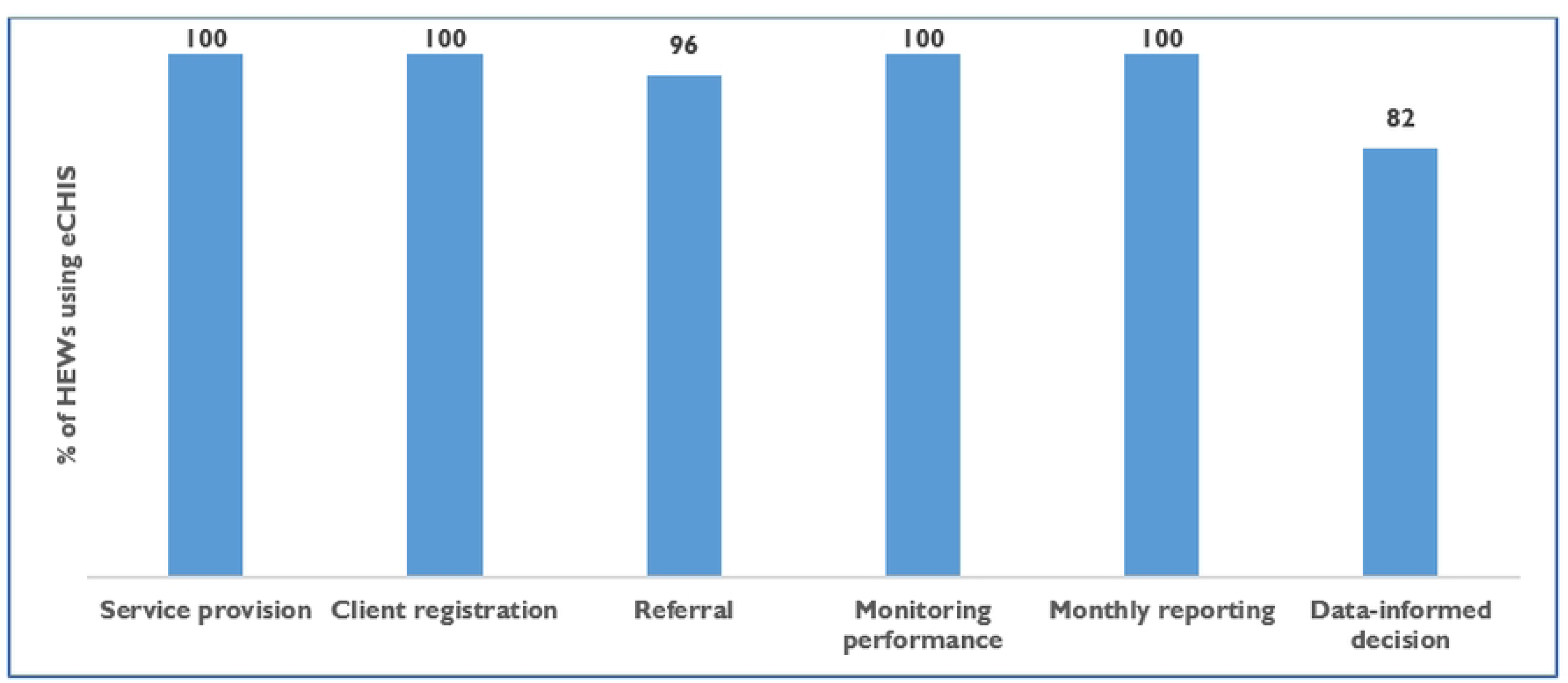
Percentage of HEWs using the eCHIS for various purposes.

Similarly, the supervisors adopted the FPA to establish their performance targets, and targets for the HEWs and the HPs on selected KPIs; provide mentorship support; conduct regular supportive supervision; monitor and track HEWs’ progress; get real-time data during performance reviews and for data-informed decisions. Twenty percent of the supervisors used FPA during integrated supportive supervision and 63% (135 of the 213) of the mentorship support was undertaken using the FPA. The extraction of raw and analyzed data from the national eCHIS dashboard was reported to be challenging for managers and supervisors. A Woreda expert said that “Extracting data from the digital platform looks easy, but it still requires skills to do so”. (KI 3).

Reviewing PBI intervention became a standing agenda during the monthly health center-level review meetings involving all health posts and HEWs in the catchment. The implementation strategies for the digitally enabled PBI innovation, which aligned with the existing health system and functions, facilitated easy adoption. The PBI innovation also assists implementers in gaining valuable insights to anticipate and promptly address bottlenecks related to essential drugs, supplies, and materials. A health center director from Mirab Abaya Woreda said “PBI helped us see how to address challenges, especially through the QI approach and root cause analysis.” (KI 4).

Challenges affecting the adoption of the intervention were related to tablet performance capacity, inconsistent internet connectivity, inconsistent power source for charging tablets, inconsistent mentorship supports, and staff turnover. A health extension worker from Mirab Abaya expressed her frustration and optimism as follows; “We achieved what we can achieve so far with the poor tablets and weak infrastructure. We will record more success in the future with improvement tablet capacity…. and the high motivation we currently have.” (HEW 7)

The eCHIS dashboard is perceived by decision-makers as a valuable tool for real-time monitoring of performance gaps. It enables them to identify areas needing improvement promptly and provide the necessary support to enhance performance effectively. As noted by a key informant from Mirab Abaya, ease of access to the real-time data helped them to see gaps. He stated, “Currently, HEWs’ performance data can be accessed in real-time from our offices. The system also enables us to identify gaps outside of work hours when necessary and guides HEWs on addressing areas of underperformance.” (KI 9)

#### Implementation

##### a) Acceptability of the innovation and implementation strategies

The acceptability of the digitally enabled PBI intervention and implementation strategies was reported as high by HEWs, supervisors, and Woreda focal persons. They appreciated the digital tools, eCHIS and FPA. The digitally enabled PBI intervention and its implementation strategies were well received by stakeholders including HEWs, supervisors, and Woreda experts. The HEWs, frontline implementers of the intervention had a positive attitude towards the digitally enabled PBI intervention for its transparency in ensuring clarity of performance targets, standards, evaluation, PBI scoring, and identification of award winners. A HEW at Awabel Woreda said, “The system ensures that those who work hard receive the incentives, and I am grateful to have been one of the recipients because I worked hard “. (HEW 1)

According to respondents, the eCHIS application and the FPA have brought convenience, simplicity, and digitalization to the workflow and HEP in general. These tools have streamlined activities, reduced manual tasks, and avoided paper-based documentation. They also stressed how these digital tools have fostered transparency and efficiency in their work, reflecting their enthusiasm and dedication to the initiatives. The FPA was reported as supportive to the supervisors in target setting, task management, and performance monitoring through the inbuilt dashboard, resulting in marked improvements in data quality and accuracy. It was also indicated that the intervention was highly beneficial in evaluating the performance of HEWs, HPs, and HCs and was described as “easily implementable”, promoting its acceptability. This was well explained by a key informant from Mirab Abaya who stated, “We accepted and implemented the digital performance management interventions with full commitment.” (KI 1). After the award, a HEW from Awabel Woreda expressed her feelings, stating, “The transparency and accuracy of the process were commendable, as it allowed me to emerge victorious, rewarding my hard work”. (HEW 1). Recognizing that their efforts would be acknowledged and rewarded, PBI intervention motivated HEWs and supervisors to work harder and perform better. The intervention’s transparency, clear KPIs, measurable criteria, and performance targets from the outset, and reliance on real-time digital data, (which is less prone to error and manipulation), contributed to trust in the system. As mentioned by a supervisor from Awabel Woreda, “Digital evaluation is an important part of PBI implementation because digital data are trustworthy, accessible, and the verification is easy.” (KI 2)

A program manager from a health center expressed performance-based incentives intervention as focused and simple, emphasizing the relevance of the strategies and careful selection of KPIs which resulted in improved service provision. The acceptability of the intervention was accepted not only because of its implementation simplicity, but also the competitive environment it created, and the winners’ spirit it fostered. The key informants emphasized the value of this spirit and its importance in boosting the morale of the staff. They also mentioned that the PBI intervention came at the right time when the HEWs were highly demotivated. The supervisors appreciated the fact that they could monitor the progress of each HC, supervisor, and HP from the digital platform. They also recognized the capacity that training and mentorship provided to HEWs allowing them to track their performance, deliver better service, and easily produce their regular reports.

A supervisor from a health center remarked, “The digital system allows us to keep track of everyone’s progress and ensures that we can offer the right support at the right time.” (KI 3). The ability to virtually oversee each HEW’s activities was a significant advancement in enhancing service delivery and providing timely remote support and feedback.

Despite its high acceptability, the implementation faced challenges due to shortages of supplies and tablets, which affected HEW’s motivation and progress. These issues were later addressed through training, supportive supervision, troubleshooting, and mentorship, improving implementation fidelity. To further enhance motivation, the Woreda health office and project staff created a Telegram channel to track performance, provide timely support, and share regular updates and feedback which fostered a sense of competition among the HEWs and HPs. A focal person from Gimbichu Woreda remarked, “HEWs, despite being heavily burdened and occupied, have shown commitment and dedication to implement the PBI interventions.” (KI 3).

##### b) Feasibility of the interventions and implementation strategies

According to health extension workers, their supervisors, the HC, and Woreda program managers, the implementation of digitally enabled performance-based incentives and their strategies was feasible. Implementing digitally enabled PBI and its strategies was feasible. Incentivizing individual HEWs and supervisors would encourage them to strive for better performance and is an effective strategy for motivating them.

The intervention was widely regarded as feasible by stakeholders, largely due to its seamless alignment with existing HEP workflow and the provision of essential resources such as tablets, Wi-Fi routers, and capacity-building training. These tools not only supported the routine operation of the system but also enhanced the efficiency and effectiveness of the performance management process. According to a HEW from Mirab Abaya, setting annual targets is a standard practice within the primary health care unit (PHCU), and the PBI intervention improved this process by introducing digital tools to streamline target-setting and performance monitoring. Further, she shared, “We filled the KPIs with my supervisors, set annual performance target breaking it down into quarters. It enables me to track my performance. I can simply compare my performance against my plan to see what I’ve achieved and what I am left with”. (HEW 8). This reflection illustrates how the PBI intervention not only aligns with existing practices but also empowers HEWs to take a more active role in tracking and managing their performance. By integrating digital tools into routine workflows, the intervention reduces administrative burdens, and supports evidence-based decision-making, thereby enhancing individual accountability and the overall functionality of the health system.

The feasibility of the PBI system was also reflected in discussions about the types and design of incentives. Financial incentives alone were the least favored by the HEWs and their supervisors. Respondents generally preferred In-kind incentives or a combination of financial, and in-kind incentives, emphasizing the long-term impact and practicality of the latter. As one HEW noted, “Money is expendable while in-kind incentive has memories that can last longer.” (HEW 2). The type of incentive provided should be tailored to the specific needs of the health posts.

Tailoring incentives to the specific needs of HPs was considered essential for ensuring their relevance and effectiveness. Informants suggested that contextually meaningful incentives, addressing felt needs and critical gaps, would not only support the health posts but also serve as a lasting acknowledgment of HEW efforts. For example, incentives focused on equipment maintenance or solar power technology were seen as more impactful than generic financial rewards. A HEW from Mirab Abaya highlighted this perspective, “If the incentives are based on the specific needs of the HP, it would be much appreciated. For example, some HPs require maintenance of their materials such as solar power, etc. The maintenance is something tangible and can be seen easily, it witnesses not only the good performances of the HEWs but also the support provided by the partners. (HEW 2). Moreover, from the perspective of institutionalizing the PBI intervention, non-financial incentives were seen as more feasible and sustainable, as they could be implemented within the health system’s existing resources and capacity. This approach aligns with the broader goal of integrating PBI into routine health systems operations.

However, there were mixed opinions on individual versus team-based incentives. While individual incentives effectively motivate HEWs and foster competition, they often do not translate into improved performance at the health post level and sometimes undermine teamwork. HEWs to be competitive, did not appear to drive improvements at the health post level as a team and often undermined teamwork. This highlights the importance of designing an incentive system that balances individual recognition with the promotion of collective effort and outcomes, further contributing to the feasibility of the PBI system. (Table 2)

**Table 2:** HEWs’ and their supervisors’ preference for incentive schemes.

| Reasons for choosing the incentive scheme | Individual | Group/team | Both |
| --- | --- | --- | --- |
| It gives recognition to individual contribution | 35% | 9% | 18% |
| It encourages individual HEWs to work hard | 62% | 36% | 35% |
| It would create positive competition among | 39% | 0% | 53% |
| It would strengthen teamwork | 23% | 64% | 47% |
| It promotes HP's performance | 31% | 18% | 53% |
| It contributes most to HEP service provision | 15% | 0% | 55% |

NB: the percentages were computed from multiple response questions which do not add up to 100%.

Through the combined approach, we can strike a balance in motivating individual effort while at the same time promoting collaboration and collective goal attainment. A HEW from Gumbichu Woreda noted: “… to me, it is better to provide incentives at the HP/team level than for the individual HEW. We are working in the same village and our efforts are similar. So, it is better to focus on the HP. For instance, if a mother comes for FP service, it creates a spirit of disruptive competition between HEWs to record the woman on the eCHIS/tablet to increase the number of provided services. The other thing is that those who don’t need the incentive might also ignore the program activities. One might say, let the one who received the incentive do all the tasks, which would affect the performance of the HP and the HEP.” (HEW 3).

Contextual factors such as population density, topography, ICT infrastructures, and HEW engagement in non-health activities contribute to performance variability among HEWs and service uptake by the community, which made the HEWs question the fairness of the performance standards, requiring thorough considerations to account for these factors in selecting KPIs and designing evaluation criteria. A HEW from a hard-to-reach health post stated “We (HEWs) were discussing that unless a geographic stratification-based incentive mechanism is implemented it is hard for us (the HEWs in hard-to-reach areas) to compete with the HPs having favorable topography… the HEWs assigned in difficult geographic locations have lots of ups and downs to serve the community. The incentive should be based on strata; highlanders with highlanders and lowlanders with lowlanders. We (the highlanders) often work long hours but can cover a few households but the lowlanders can reach many households with a comparable given time.” (HEW 3).

Similarly, denominator issues for reliable target setting have been issued by HEWs and their supervisors on the target population/denominator for setting performance targets. Participants challenged the validity of the woreda-prescribed target for the KPIs as there were variations in the target population when the actual counted population was considered. During a discussion at the FGD, with HC supervisors, they expressed, “The annual performance target given by the woreda is not in line with the reality on the ground. This also includes the way the conversion factor is designed and developed. The conversion factor is high. Though we understand the challenges, we have no way out other than using the prescribed conversion factor and population estimate.” (FGD 2). Similar concerns were raised during FGD with HEWs at Awabel Woreda, who emphasized the importance of using the actual number of populations instead of Woreda’s projected population. They stated as follows, “Setting KPIs target should be based on the actual Kebele counted population. The target set at the woreda level has problem … but the woreda does not trust the eCHIS counted population.” (FGD 3)

#### Maintenance

The study used qualitative data to assess the sustainability of PBI interventions and the influencing factors. Mixed perspectives emerged. On one hand, sustainability seems achievable, as the KPIs align with the routine health system’s tracking mechanisms, and most implementation strategies are already integrated into the existing support and performance management systems of the HEP and community health structures. As a result, sustaining the intervention is unlikely to pose significant challenges for the community health system. A key informant from the Mirab Abaya Woreda health office expressed, “We don’t consider the implementation as an extra task. It assisted us in accessing the data easily and documenting service delivery in a quality way. The PBI KPIs are routinely planned HEP services that are being performed and are not new at all.” (KI 10). On the other hand, the sustainability of the eCHIS faces significant challenges due to a lack of dedicated resources, budget allocation, and a defined accountability structure. These gaps hinder the ability to sustain key components such as regular mentorship and incentivization, which are currently not integrated into the routine support system. A HEW from Mirab Abaya expressed this as: “The government’s current status can’t let it organize training, incentives and provide required inputs. Allocation of budget for the program should precede before the handover.” (HEW 6). A similar reflection from a key informant at Mirab Abaya Woreda stated, “… My fear is the continuity of the PBI when fully implemented by the government. Clear guidance and accountability of the eCHIS should be in place under the structure of the health system to use the budget. So far, we are implementing the PBI strategies (like the mentorship and review meetings) through integration with other health system activities using other programs’ budgets… unavailability of dedicated finance from the government would be a challenge for the continuity of mentorship and required support for the PBI. The eCHIS program has not been appropriately placed in the Woreda hierarchical structure. I have concerns about its continuity without it being budgeted.” (KI 10). These reflections underscore the need for proper financial planning, a defined program structure, and accountability mechanisms within the health system to ensure the sustainability and effectiveness of the eCHIS program.

The informants emphasized the need for JSI to provide support until the health system or government fully assumes ownership of the intervention by establishing an accountable structure and allocating the necessary budget and resources. This was reflected by a HEW from Mirab Abaya who stated “The partner should work until the government starts allocating the budget to it. Continuity of logistics provision related to the tablet and other items should be there to ensure the running of the eCHIS. Apart from this HEP services provision is the routine work we are paid for.” (HEW 6)

Similarly, a Key informant from the Mirab Abaya Woreda health office highlighted the critical role of ensuring uninterrupted supply and maintenance of key tools like tablets, comparing them to essential service-providing equipment such as blood pressure apparatus and weight scales. He remarked: “The tablet is now like a service-providing tool, the same as blood pressure apparatus, weight scale, etc.; so, the government should uninterruptedly supply it in case of damage, and maintenance should also be well planned before the partner handover it to the government. The absence of these supplies after the full-scale implementation means allowing the health extension program to collapse at some point. ‘’ … the solution to this is proper budgeting for the eCHIS program” (KI 10).

## Discussion

The digitally enabled performance-based incentive intervention demonstrated high acceptability, feasibility, and effectiveness, significantly enhancing HEWs’ motivation and performance while improving RMNCH outcomes. Supervisors effectively used digital tools to set performance targets, while eCHIS adoption enabled real-time tracking, streamlined workflows, and data-driven decision-making. However, the sustainability of the PBI intervention, though supported by its alignment with routine health systems, is constrained by the absence of dedicated resources, budgets, and institutional support within the government health system. Key findings from this implementation research underscore the significance of digitally enabled performance-based incentives (PBIs) in transforming the performance management system for HEP. The participatory design process with extensive stakeholder engagement was significant for the intervention’s success. This approach ensured the contextual relevance and cultural acceptability of the interventions, fostering a sense of ownership among stakeholders, easy adoption for the intervention, and feasibility for implementing it within the HEP. Respondents appreciated the digital tools that minimized human intervention in performance monitoring, enhancing trust and reliability, and their contribution to improving motivation and RMNCH outcomes. The introduction of the FPA and eCHIS dashboard, with features for target setting, supervisory tasks, and dashboards, was pivotal in streamlining operations and facilitating the acceptance of the PBI interventions.

These results align with global evidence that supports the use of digital innovations to strengthen community health systems, particularly in resource-limited settings [20].

The FPA’s automation of supervisory functions proved instrumental in improving HEWs’ performance. The functionalities offered by the FPA streamline and improve various operational and performance management functions contributing to the overall acceptance and effectiveness of the PBI innovation. According to Agarwal S, Sripad P, Johnson C, Kirk K, Bellows B, Ana J, et al. [21], digitized health information systems brought opportunities to address the long-standing challenge of measuring community health workforce performance (18). Employing digital tools that minimize human manipulation in monitoring and measuring KPIs fosters a sense of trust in the system. Consistent with our findings, a study has shown the significance of digital health tools in improving operational programs efficiency facilitating improved planning, supervision, and performance monitoring [10]. By providing real-time data for tracking and personalized feedback, the application enabled supervisors to identify and address performance gaps more effectively. This streamlined approach to supervision reduced administrative burdens and allowed for a greater focus on mentoring and capacity building. The alignment of CHW activities with national health priorities through target-setting further enhanced accountability and service quality (Mettler and R.P. 2009). These findings underscore the importance of integrating technology into supervisory frameworks to optimize health service delivery, acceptability, and intervention adoption. The integration of PBIs with digital tools provided a scalable and sustainable model for enhancing accountability and performance within HEP. These findings are consistent with global studies demonstrating the effectiveness of PBIs in motivating health workers and improving service utilization [22].

The adoption of digitally enabled PBI was evident in its full integration in all intervention health posts, health centers, and Woredas reflecting the success of PBI integration into the existing system and HEP workflow. The HEW extensively uses the eCHIS for population and household registration, HEP service provision, client referral, tracking progress, generating monthly reports, and extracting and using the data for making informed decisions at the local level. The supervisors use the FPA for performance target setting, monitoring HEW performance, conducting supportive supervision and mentorship, and using the data from the dashboard for reporting and program management. The systematic use of digital tools enhanced accountability, streamlined support and reporting processes, and effective service delivery. The informants consistently highlighted the system’s alignment with existing health structures as a key factor for supporting adoption. Implementing stakeholders found the PBI framework feasible and adaptable to local contexts. HEWs reported that the system simplified target tracking and reduced administrative burdens. The intervention’s feasibility was reinforced by its alignment with existing workflows and its capacity to address identified gaps in the health system.

Challenges with infrastructure such as unreliable connectivity and power supply, internet connectivity, and electric power supply to charge the tablets can affect PBI intervention. These factors must be carefully considered to ensure the successful institutionalization of the intervention, and adoption of the technology as they can hinder effective implementation and usage [10], [23] Respondents noted that digitalization fostered a sense of fairness in performance evaluations and awards. Despite its positive contribution in instilling competitiveness, individual-level awards were concerning among implementing stakeholders as they would damage teamwork and be less significant in driving HEP goal attainment. Balancing individual motivation with collective goals fosters a collaborative environment among HEWs, reinforcing the potential for performance-based incentives (PBI) to enhance overall performance while maintaining team cohesion [24]. This hybrid model successfully drove performance improvements at individual and health post levels without negatively impacting collaboration. According to a study conducted by Martin B, Jones J, Miller M, and Johnson-Koenke R. (2020) [24], good performance was associated with intervention designs involving a mix of incentives, frequent supervision, continuous training, community involvement, and strong coordination and communication between CHWs and health professionals, leading to increased credibility of CHWs. The HEWs preferred recognition at the health post level rather than as individuals, as they work closely together in the same community, a focus on team performance rather than individual is more meaningful. It is also aligning with the collective nature of healthcare delivery indicating that a well-structured PBI system can effectively motivate teams.

Preferences for mixed financial and non-financial incentives emerged, reflecting the importance of balancing tangible rewards with lasting benefits. According to Cok, 2015, [7], mixing financial and non-financial incentives can be an effective strategy to enhance performance, when they are predictable and communicated to potential implementers. The preferred incentive systems align with the existing less structured, manual, and often biased incentivizing mechanisms implemented by the health system sporadically. Implementing non-financial incentives is more feasible for HEP than financial from the perspective of accounting and resource allocation. The allocation of adequate public funding is essential for the successful implementation of health initiatives, including incentivization mechanisms [3]. By aligning financial resources with health priorities and outcomes, governments can create a more stable environment for implementing such programs. Variability in geographic and infrastructural conditions prompted calls for stratified incentive models to ensure equity.

Recognizing these contextual factors helps develop equitable performance standards to foster a sense of ownership and commitment among HEWs, which further enhances, the acceptability, adoption, and feasibility of implementing PBIs tailored to local conditions [25].

Incentivization was a key driver of improved motivation among HEWs, with qualitative accounts emphasizing the role of transparency and competition. Incremental improvements in KPI scores during subsequent PBI award rounds highlighted the intervention’s role in fostering sustained motivation and performance enhancements.

To create enabling conditions for implementing the intervention HEWs, supervisors and Woreda managers received training and essential supplies. The PBI and FPA training effectively equipped implementing stakeholders with the knowledge and skills needed to implement digitally enabled PBI, evidenced by significant improvement in knowledge and skills scores post-training and positive reactions to the training. Digital tools were distributed to all targeted HEWs and supervisors, facilitating comprehensive engagement. Intervention designs that involved frequent supervision and continuous training led to better CHW performance in certain settings [7], [22].

The intervention significantly enhanced maternal and child health service provision, as evidenced by increases in eCHIS-recorded ANC visits and immunization rates. Time-series analyses demonstrated consistent performance improvements in intervention Woredas. For instance, the average ANC first visit rate tripled post-intervention, contrasting with a decline in comparison areas. Similarly, growth monitoring indicators showed upward trends in intervention regions, underscoring the intervention’s positive impact on service delivery outcomes.

## Limitations

The study faced limitations that impacted its implementation and outcomes. Revisions in administrative boundaries disrupted support for some health posts, while national campaigns diverted HEWs from routine activities. Insecurity in certain regions, unreliable internet connectivity, inconsistent electricity supply, and inadequate availability of essential drugs and supplies further constrained operations. Challenges with the maturity of the eCHIS system, data quality, report consistency, and dashboard usability were notable. Limited financial resources, turnover of trained HEWs and supervisors, competing priorities, and inconsistent mentorship and supervisory support hindered effective implementation. Contextual variations in population size, topography, HEW density, and infrastructure also complicate performance management and target setting.

## Conclusion

The performance-based incentive innovation was proven to be feasible and effective in the Ethiopian HEP. Feasibility was demonstrated by its integration into existing health system workflows and operations and alignment with existing practices. The provision of digital tools (tablets, Wi-Fi routers, power banks) made implementation smooth, and training and capacity building enabled HEWs and their supervisors to use the tools.

HEW supervisors used the Enhanced Focal Person Application to set targets, track progress, and provide digital supervision and mentorship support. However, infrastructure gaps (poor internet, inconsistent electricity, limited tablet performance) hindered full implementation. Inconsistent mentorship and high staff turnover were also challenges. While the innovation was short-term feasible, long-term sustainability requires dedicated resources, institutional ownership, and proper budget allocation.

The intervention showed a high impact on HEW performance, motivation, and maternal and child health outcomes. HEWs showed significant improvement in KPI scores, with an average score of 66 to 82 during the second round (p<0.05). Supervisors and managers observed increased motivation and accountability among HEWs due to clear and measurable incentives and a transparent performance evaluation process. Service delivery outcomes improved significantly, with the average number of ANC first visits tripling (from 56 to 155, p<0.001) and significant gains in childhood immunization and growth monitoring indicators. Real-time data from the eCHIS dashboard enabled data-driven decision-making, monitoring, supervision, and service delivery. Stakeholders praised the system for being fair, and transparent and building trust through digital tools.

Despite the achievements, infrastructure and geographic variation in performance evaluation sometimes limited the impact. Overcoming these limitations and ensuring equitable implementation across different contexts will be crucial. Overall, the PBI is proven to strengthen community health systems, service delivery, and accountability, a model that can be scaled up in other low-resource settings.

The long-term impacts of digitally enabled PBIs on health outcomes, workforce motivation, and systemic efficiency call for further evaluations. These lessons provide a roadmap for strengthening community health systems globally, with the potential to improve health equity and service delivery in low- and middle-income countries.

## Data Availability

Data will be available on eCHIS CommCare and National eCHIS Dashboard for analysis. For qualitative data, audio recordings are also available for reference

https://echis-dash.moh.gov.et/dashboard/list/?pageIndex=0&sortColumn=changed_on_delta_humanized&sortOrder=desc&viewMode=table

## Acknowledgements

We are indebted to the Ministry of Health, and other stakeholders for their invaluable support for the realization of this manuscript. Our sincere thank also goes to the communities, data collectors, and support teams whose contribution have been indispensable for the success of this work.

